# Robustness Gap of Large Language Models in Nephrology

**DOI:** 10.64898/2026.08.17.26360565

**Authors:** Ayaka Soejima, Fumiya Kitano, Daisuke Ichikawa, Yugo Shibagaki, Ryunosuke Noda

**Affiliations:** Division of Nephrology and Hypertension, Department of Internal Medicine, St. Marianna University School of Medicine, 2-16-1, Sugao, Miyamae-Ku, Kawasaki, Kanagawa, 216-8511, Japan

**Keywords:** Artificial intelligence, Clinical reasoning, Nephrology, Large language model, ChatGPT

## Abstract

**Background:** Whether benchmark performance reflects robust clinical reasoning rather than surface-level pattern recognition remains uncertain. We evaluated the robustness of state-of-the-art large language models (LLMs) on nephrology board renewal questions using “None of the other answers” (NOTA) substitution.

**Methods:** From 210 Japanese Society of Nephrology board renewal questions (2014–2023), two nephrologists independently reviewed all items. Questions in which NOTA became the sole correct answer after replacement were included, yielding 145 validated questions. GPT-5, GPT-4o, Gemini 2.5 Pro, and Gemini 2.0 Flash were evaluated via application programming interfaces under default settings. The primary endpoint was accuracy, and paired differences were assessed using the exact two-sided McNemar test.

**Results:** Accuracy was significantly lower after NOTA substitution for all models: GPT-4o, 66.21% to 19.31% (drop, 46.90 percentage points [pp]); GPT-5, 87.59% to 73.10% (14.48 pp); Gemini 2.0 Flash, 58.62% to 31.03% (27.59 pp); and Gemini 2.5 Pro, 86.90% to 55.86% (31.03 pp); all P < .001. GPT-5 showed the smallest decline and the highest accuracy in both versions.

**Conclusions:** All evaluated LLMs showed a significant robustness gap after NOTA replacement. Newer models may be more robust, but multiple-choice accuracy remains an incomplete measure of clinical reasoning robustness.

---

Large language models (LLMs) have rapidly advanced and are increasingly explored for clinical support. In nephrology, GPT-4 achieved a total accuracy of 74% on multiple-choice, single-answer questions from nephrology and kidney self-assessment programs, but still performed below passing thresholds and the average score of nephrology examinees [1]. o1 pro has also achieved passing scores on nephrology board renewal questions, further heightening expectations for real-world use [2].

However, autonomous clinical deployment—where AI systems independently finalize diagnoses and treatments without human oversight—remains premature. Because LLMs can still generate unpredictable errors in complex clinical scenarios, they lack the reliability required for critical medical decision-making. Consequently, current explorations in nephrology focus primarily on safer, supportive applications like patient education and dietary counseling [3].

A central concern is whether benchmark performance reflects robust clinical reasoning rather than surface-level pattern recognition. Across medical settings, LLM performance can drop when tasks shift from multiple-choice to free-response or multi-turn patient-interaction formats that better approximate clinical reasoning demands [4]. Therefore, evaluation strategies that probe robustness—not only accuracy—are increasingly important.

One simple robustness stress test is to replace the original correct option in a multiple-choice question with “None of the other answers” (NOTA), forcing models to reject all remaining options if they truly follow the underlying reasoning [5]. A recent study applying this approach to a general medical benchmark reported substantial accuracy declines across leading LLMs [6]. While general medical questions can sometimes be resolved through superficial keyword associations, nephrology demands complex clinical reasoning—such as the deep integration of laboratory findings and pathophysiology required to determine kidney biopsy indications. Therefore, whether a similar robustness gap exists in such highly specialized nephrology scenarios remains to be elucidated.

Here, we applied NOTA substitution to nephrology board renewal questions and evaluated state-of-the-art LLMs from OpenAI and Google. We hypothesized that models capable of genuine causal, pathophysiology-based reasoning would be less affected than those relying on shortcut learning via answer-pattern matching.

From 210 Japanese Society of Nephrology board renewal questions (2014–2023), two nephrologists independently reviewed all items. Only questions in which NOTA would be the sole correct answer after replacement were included, with discrepancies resolved by consensus, resulting in a final set of 145 validated questions. Four LLMs—GPT-5, GPT-4o, Gemini 2.5 Pro, and Gemini 2.0 Flash—were evaluated via their application programming interfaces under default settings. Each question pair (original and NOTA-modified) was presented independently, except for sequential questions from the same case, which were grouped to maintain clinical context. The primary endpoint was the proportion of correct answers. Significance was assessed using the exact two-sided McNemar test, and 95% confidence intervals (CIs) for accuracy drops were calculated via 1,000-iteration bootstrapping (P < .05 was considered statistically significant). Analyses were performed using Python.

Across all 145 questions, accuracy on the NOTA-modified version was significantly lower than on the original version for all four models (Table 1). GPT-4o accuracy dropped from 66.21% (96/145) to 19.31% (28/145), a decrease of 46.90 percentage points (pp) [95% CI, 37.93–56.55; P < .001]. GPT-5 declined from 87.59% (127/145) to 73.10% (106/145), a drop of 14.48 pp [95% CI, 8.28–21.38; P < .001]. Gemini 2.0 Flash decreased from 58.62% (85/145) to 31.03% (45/145), a drop of 27.59 pp [95% CI, 17.93–36.55; P < .001]. Gemini 2.5 Pro fell from 86.90% (126/145) to 55.86% (81/145), a decline of 31.03 pp [95% CI, 22.76–39.31; P < .001]. Among the four models, GPT-5 showed the smallest decrease and achieved the highest accuracy in both versions, though its decline remained statistically significant.

**Table 1.**
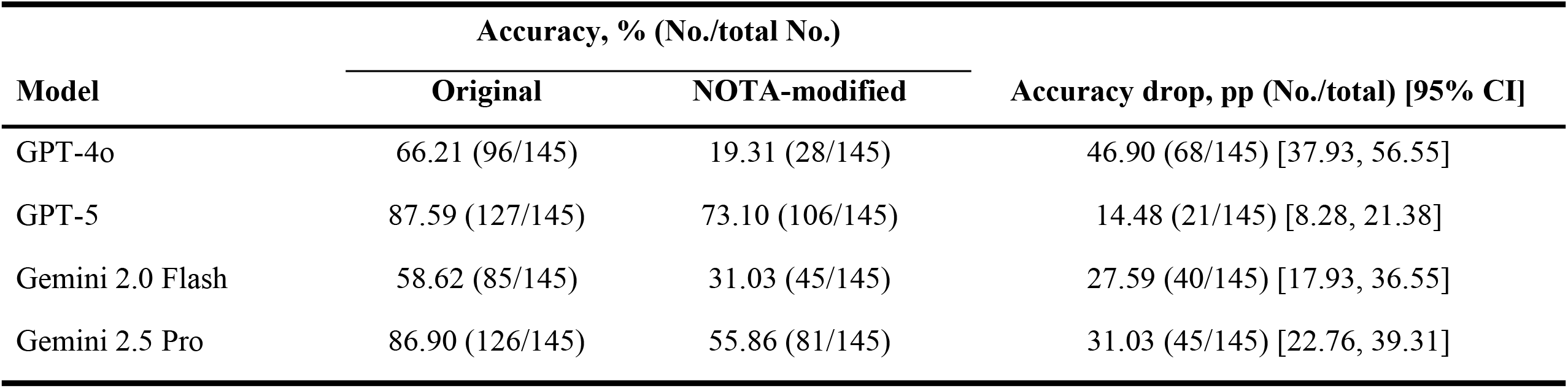
Performance on original and NOTA-modified questions.

| Model | Accuracy, % (No./total No.) |  | Accuracy drop, pp (No./total) [95% CI] |
| --- | --- | --- | --- |
|  | Original | NOTA-modified |  |
| GPT-4o | 66.21 (96/145) | 19.31 (28/145) | 46.90 (68/145) [37.93, 56.55] |
| GPT-5 | 87.59 (127/145) | 73.10 (106/145) | 14.48 (21/145) [8.28, 21.38] |
| Gemini 2.0 Flash | 58.62 (85/145) | 31.03 (45/145) | 27.59 (40/145) [17.93, 36.55] |
| Gemini 2.5 Pro | 86.90 (126/145) | 55.86 (81/145) | 31.03 (45/145) [22.76, 39.31] |

Overall, all evaluated LLMs showed statistically significant accuracy declines after NOTA replacement. GPT-5 achieved the highest accuracy in both conditions and exhibited the smallest decrease. These findings suggest that newer models may be more robust, but multiple-choice accuracy remains an incomplete measure of reasoning robustness.

Our results extend prior NOTA-based observations from a general medical benchmark to a nephrology-specific, Japanese-language question set [6]. They also align with evidence that medical LLMs can be brittle under other clinically relevant perturbations, including multi-turn follow-up interactions [7] and terminology shifts such as drug-name substitutions [8]. Taken together, these data suggest that robustness gaps may reflect a broader vulnerability of current LLMs rather than a dataset- or language-specific artifact.

One plausible explanation is that LLMs partially anchor on answer-choice priors: when the original correct option is removed, models may select the most familiar or semantically proximate distractor instead of fully eliminating all remaining choices to reach NOTA. This interpretation is consistent with recent evidence that LLMs often fail to recognize unanswerable medical questions or absent correct options, even when they respond with high confidence [9]. Clinically, the concern is similar: when essential information is missing, an LLM may still choose a plausible answer rather than state that the case cannot be judged reliably.

This issue is particularly relevant to nephrology, where clinical decisions often depend on integrating laboratory values, longitudinal trends, and physical findings. In real-world practice, LLMs may not have access to a complete medical record, and important contextual information—such as subtle physical examination findings regarding volume status—may be absent or difficult to encode in text prompts. A safe model should recognize such information gaps and request additional data or defer judgment. Instead, the behavior observed in NOTA testing raises the concern that models may overcommit to a plausible but incorrect management pathway based on fragmented inputs. Such input-quality dependence has also been identified as a limitation of current LLM applications in nephrology [3].

This study has limitations. NOTA substitution is an artificial manipulation and may not directly quantify real-world clinical safety. All questions and responses were in Japanese, and generalizability to other languages is uncertain. The test set reflected general nephrology, and robustness may vary across subspecialties. Finally, performance may differ under alternative prompting strategies or system configurations. Despite these limitations, the consistent NOTA-related accuracy loss observed across major LLMs supports routine reporting of robustness metrics alongside standard accuracy when considering nephrology applications.

## Author contributions

Conceptualisation: A.S., R.N.; Methodology: R.N.; Data curation: A.S., R.N.; Formal analysis: R.N.; Writing—original draft: A.S.; Writing—review & editing: all authors; Supervision: F.K., D.I., Y.S., R.N.

## Acknowledgements

The results of this study have not been published previously in full or in part.

## Data availability

The source question set is proprietary to the Japanese Society of Nephrology and cannot be publicly shared. De-identified evaluation outputs and analysis code are available from the corresponding author upon reasonable request.

## Ethical approval

Not applicable. This study did not involve human participants, patient data, or interventions.

## Informed consent to participate

Not applicable. This study did not involve human participants, patient data, or interventions.

## References

1. Miao J, Thongprayoon C, Garcia Valencia OA, et al (2024) Performance of ChatGPT on nephrology test questions. Clin J Am Soc Nephrol 19:35–43. 10.2215/CJN.0000000000000330

2. Noda R, Yuasa C, Kitano F, Ichikawa D, Shibagaki Y (2025) Performance of o1 pro and GPT-4 in Self-Assessment Questions for Nephrology Board Renewal. Front Med (Lausanne) 12:1702668. 10.3389/fmed.2025.1702668

3. Unger Z, Soffer S, Efros O, et al (2025) Clinical applications and limitations of large language models in nephrology: a systematic review. Clin Kidney J 18:sfaf243. 10.1093/ckj/sfaf243

4. Johri S, Jeong J, Tran BA, et al (2025) An evaluation framework for clinical use of large language models in patient interaction tasks. Nat Med 31:77–86. 10.1038/s41591-024-03328-5

5. Sánchez Salido E, Gonzalo J, Marco G (2025) None of the others: a general technique to distinguish reasoning from memorization in multiple-choice LLM evaluation benchmarks. arXiv preprint arXiv:2502.12896. 10.48550/arXiv.2502.12896

6. Bedi S, Jiang Y, Chung P, et al (2025) Fidelity of medical reasoning in large language models. JAMA Netw Open 8:e2526021. 10.1001/jamanetworkopen.2025.26021

7. Manczak B, Lin E, Eiras F, O’Neill J, Mugunthan V (2025) Shallow robustness, deep vulnerabilities: multi-turn evaluation of medical LLMs. arXiv preprint arXiv:2510.12255. 10.48550/arXiv.2510.12255

8. Gallifant J, Chen S, Moreira P, et al (2024) Language models are surprisingly fragile to drug names in biomedical benchmarks. Findings of the Association for Computational Linguistics: EMNLP 2024:12448–12465. 10.18653/v1/2024.findings-emnlp.726

9. Griot M, Hemptinne C, Vanderdonckt J, Yuksel D (2025) Large language models lack essential metacognition for reliable medical reasoning. Nat Commun 16:642. 10.1038/s41467-024-55628-6

